# An extended SIR model with vaccine dynamics for SARS-CoV-2 adaptation rate

**DOI:** 10.1101/2022.02.11.22270784

**Authors:** R. Horvat, T. Surić

## Abstract

The COVID-19 pandemic is caused by infection with a SARS-CoV-2 virus, a RNA virus characterized by high mutation and replication rates. From epidemiological perspective, the trajectory in time of viral adaptation is determined by two opposite forces: (a) proportion of the population who has acquired immunity, exerting thereby a selective pressure on the virus and (b) proportion of infected people in the population, measuring the net viral load in the population. We calculate both the number of advantageous mutations in the population that have accumulated by time t and the amount of viral adaptation transmitted to a susceptible compartment, the latter being called the Evolutionary Infectivity Profile (EIP). To this end we employ first a simple compartmental SIR model with a single parameter describing reduction in transmission due to vaccination. Then we switch to a model which actuates the full vaccine dynamics, including vaccination rate and short duration of immunity. In our models we have never come across a situation where vaccination plays a dominant role in driving new SARS-CoV-2 variants. Nevertheless if the vaccination rate is not high enough, the EIP would continuously grow with time, pointing to a fruitful field for proliferation of genetically variable SARS-CoV-2 viruses.

## I. INTRODUCTION

The best control strategy for handling a situation where multiple outbreaks of infectious disease are likely to occur, as in the COVID-19 pandemic, was laid down some time ago [1]. However it has been soon noticed that even with large-scale vaccination, for a disease with such a high contagiousness and short-lived immunity, the creation of herd immunity might be elusive [2]. SARS-CoV-2, the virus responsible for COVID-19, has been subject to a rapid genetic drift since the onset of pandemic in late 2019 [3]. Another challenge would be to reckon to what extent vaccination drives the evolution of variant SARS-CoV-2 viruses. This has become especially pressing with the appearance of the Omicron variant, the second most contagious but the fastest spreading virus in the world. A relationship between virus adaptation and average immune pressure in individual hosts was sketched first in [4]. Here the two basically different forces are in action. In the main vaccination will increase population immunity, thereby increasing the strength of selection for immune escape. Inversely, vaccination will decrease the net viral load in the population, hence depleting the possibility for mutations to result in a new variant of the virus. Which of these two forces would ultimately prevail? An attempt is being made in the present paper to answer this question by considering the two epidemiological models. Incidentally, our analysis would also be relevant for any similar disease apart from COVID-19. For illustrative purposes of the problem, we begin with the simplest model possible, the one in which the basic SIR compartmental model is supplemented with a single parameter representing the fraction of vaccinated people in the population. The long-term immunity is assumed for both the recovered and the vaccinated. Next we set to a thorough analysis in which the full vaccine dynamics is taken into account, including a short duration of immunity both for the recovered and the vaccinated.

## II. DEFINITIONS

Here we adopt some basic definitions from [4], and modify them to matter epidemiologically. The number of advantageous mutations that have accumulated by time t in the population is proportional to

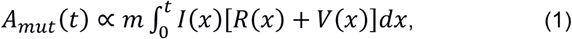

where *m* is the viral mutation rate per capita per unit time, *I(t)* is the proportion of infected/infectious people in the population at a time *t, R(t)* is the proportion of the population who had recovered from a disease and acquired immunity at a time *t* and *V(t)* is the proportion of the population who had been vaccinated and acquired immunity at a time *t*. Here the sum, *R*(*t*)*N* + *V*(*t*)*N*, where *N* is the total number of people in the population, represents a net immune response of the population at a time t. The potential for transmission of these mutations (termed as Evolutionary Infectivity Profile (*EIP*) in [4]) to the susceptible compartment *S(t)* must be proportional to *I(t)*, hence

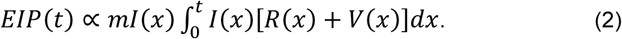

Note that *EIP(t)* weighs the average amount of viral adaptation transmitted to the susceptible compartment at a time *t* after the onset of the infection. Further, if the number of infected tends to zero for large times, the *EIP* would do the same. We shall show in the following that for an infectious disease with a short duration immunity and for a low vaccination rate, this might not be the case. This would inevitably get us into a potentially dangerous situation, in which a virus quickly changes. Trevor Bedford was the first to think, admittedly heuristically, about this problem for the SARS-CoV-2 virus [5].

## III. A SIMPLE MODEL

To illustrate how the above formulas apply when we have both the recovered and the vaccinated in the population, we start with a simple compartmental SIR model [6] and [7]. The dynamics of susceptibles *S*, infecteds *I* and recovereds *R* is described by the following differential equations:

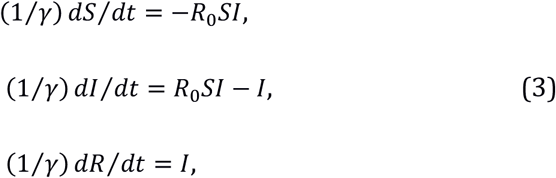

where *γ* is the rate of recovery from infection and *R*_*0*_ is the basic reproduction number. The initial conditions at the onset of the infection are *I*(0) ≠ 0, *R*(0) = 0 and *S*(0) = 1 − *I*(0) − *f*, where the parameter *f* represents the reduction in transmission due to the presence of the fraction *f* of vaccinated in the population at the onset of the infection. The effective reproduction number *R*_*eff*_ *= (1-f) R*_*0*_ is established at *t=0* in our simplistic model. Note that as the stock of vaccinated people stays constant over time, it does not enter the equations, but should be present kinematically in order to express the constancy of population N at any moment. Formally, for later use in a more general discussion, we may introduce the compartment of vaccinated *V(t)* and set *V*(*t*) = *f*, with *dV*/*dt* = 0. For *t* → ∞, *S, I* and *R* approach their asymptotic values *I*(∞) = 0, *R*(∞) and 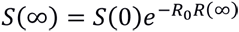, where *R*(∞) and *S*(∞) are determined from the condition 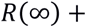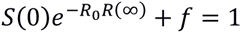, which is a simple transcendental equation for *R*(∞).

In Fig. 1 we have drawn a plot with the viral abundance ***I***(t) on the y-axis, and the population immune pressure, *R*(*t*) + *V*(*t*), on the x-axis. We shall call it the Viral Abundance - Immune Pressure (VA-IP) diagram. The hyperbolas on the plot are the curves on which the product *I*(*t*)[*R*(*t*) + *V*(*t*)] stays constant, rising from the bottom up. Note that when we switch from *f* = 0 to a finite *f*, the viral adaptation rate penetrates less and less into the curves with ever increasing values, as we increase f. Also the *EIP* switches off for large times for any *f*, as the compartment ***I***(t) eventually empties off. As *f* is increasing, the maximum of *I*(*t*)[*R*(*t*) + *V*(*t*)] shifts towards larger times, but the number of accumulated mutations always decreases. However the *EIP* shows an interesting feature (see Fig. 2). We see that curves with various *f*”s do cross each other at some instant of time. This is a consequence of the fact that the more vaccinated people are in the population, the later the maximum of infection will be attained. The potential for transmission of virus mutations at late time is thus larger in the vaccinated case. In our figures we have taken *R*_o_ = 6, a lower limit for the highly contagious and the predominant SARS COV2 variants nowadays, the Omicron. We have also taken for the infectious period 1/*γ* to be 4 days.

**Fig. 1.**
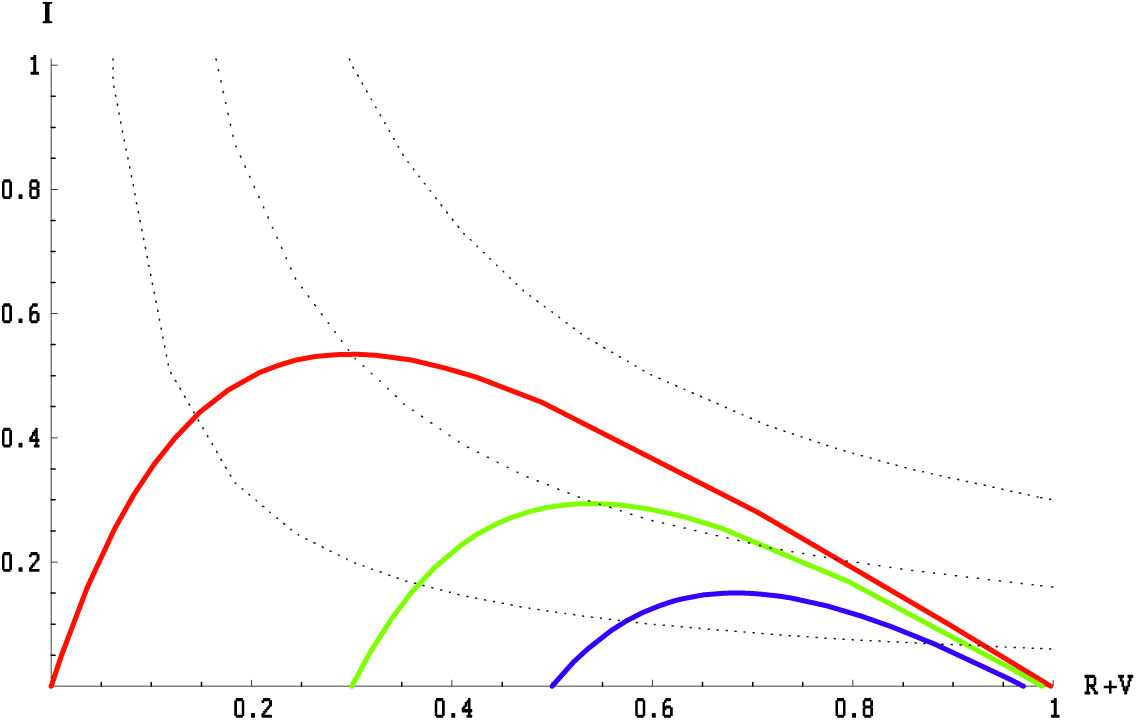
The epidemic dynamics described by the SIR model, Eq. (3), is illustrated in the VA-IP diagram with ***I***(t) on the y-axis, and the population immune pressure, *R*(*t*) + *V*(*t*), on the x-axis. The colored lines show the epidemic dynamics for *f*=0 (red line), *f*=0.3 (green line) and *f*=0.5 (blue line), *R*_o_ = 6, γ=1/4. In these cases the epidemic starts with *I*(0) = 10^−4^, *R*(0) + *V*(0) = *f* and ends with *I*(*t* → ∞) = 0. The dotted hyperbolas represent lines with the constant adaptation rate *I*(*t*)[*R*(*t*) + *V*(*t*)].

**Fig. 2.**
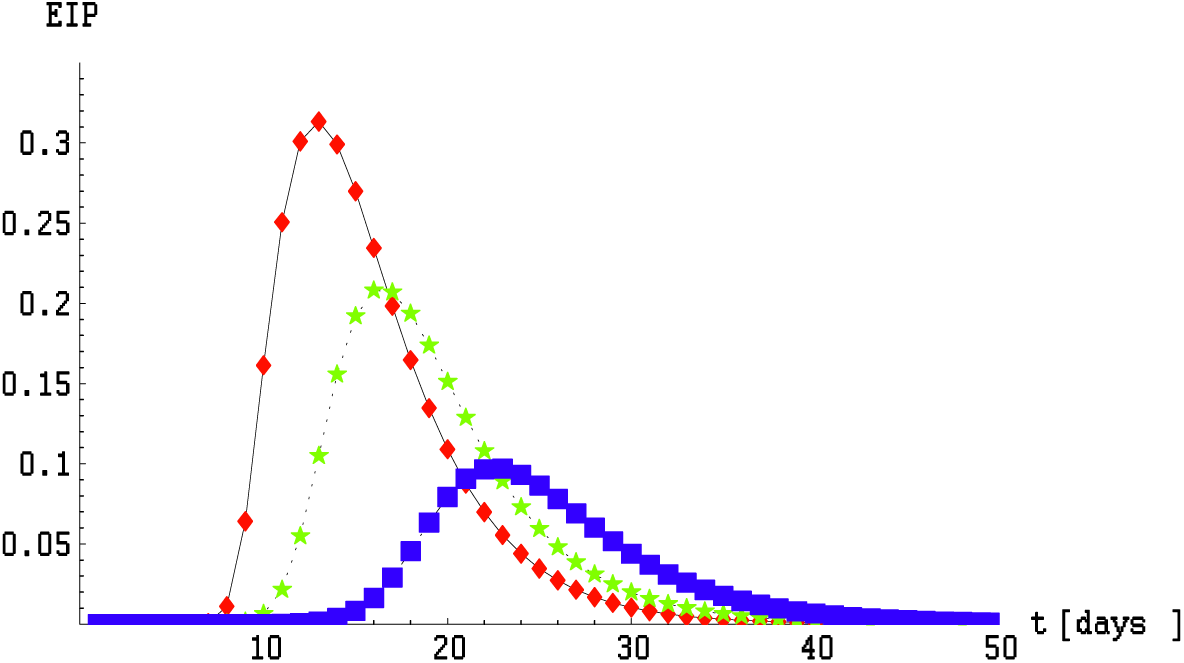
*EIP*, Eq. (2), calculated within the SIR model as a function of time for *f*=0 (red line), *f*=0.3 (green line) and *f*=0.5 (blue line), *R*_o_ = 6, γ=1/4. Initial conditions are the same as in Fig. 1.

## IV AN EXTENDED MODEL WITH VACCINE DYNAMICS AND SHORT DURATION IMMUNITY

We define our model with vaccine dynamics and short duration immunity by the following set of differential equations:

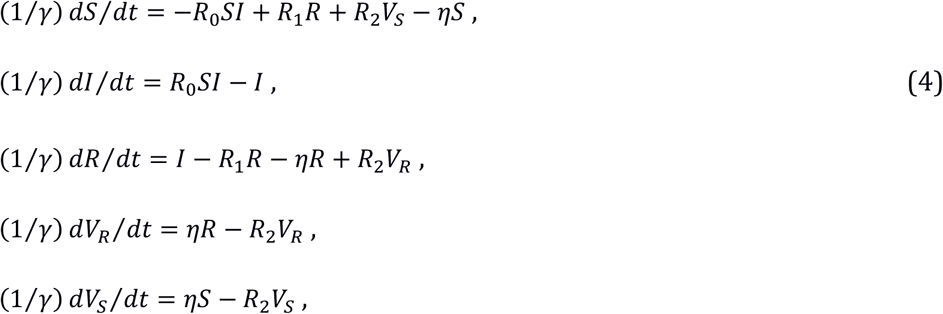

where *R*_1_ = *γ*_1_/*γ, R*_2_ = *γ*_2_/*γ, η* = *ζ*/*γ, γ*_1_ is the rate of losing immunity vested by getting over a disease, *γ*_2_ is the rate of losing immunity vested by vaccination (the rate at which those people are being vaccinated is modulated by vaccine efficacy for transmission) and *ζ* is the rate at which susceptible and recovered people acquire immunity by vaccination. The parameters *γ* and R_0_ are the same as in Eq. (3). The stock of vaccinated people splits as *V* = *V*_S_ + *V*_R_. The same *ζ* is assumed for both *V*_S_ and *V*_*R*_.

An important remark is in order at this point. There is a reason we have had to split the compartment of vaccinated individuals into the two, *V*_S_ and *V*_R_, both of which obey a separate differential equation. We could have joined the last two equations into a single one (modifying the rest of the equations accordingly),

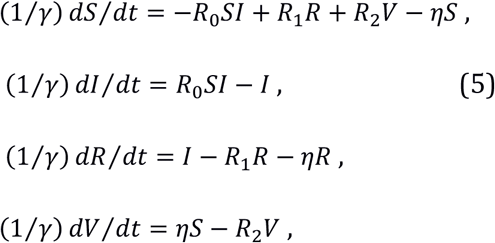

but in this case the model would be accurate only if *γ*_1_ *≥ γ*_2_. In (5), after a time 1/*γ*_1_ had elapsed, the compartment of vaccinated emptied out entirely into the compartment of susceptibles. This is altogether consistent if *γ*_1_ *≥ γ*_2_. But for *γ*_1_ < *γ*_2,_, our set of equations should provide for a possibility for the compartment of vaccinated to empty out into the compartment of recovered as well, since duration of naturally vested immunity lasts longer than duration of immunity vested by vaccination. Otherwise an individual with (for instance) a lifelong immunity (*γ*_1_ = 0) would be driven, through vaccination, again to susceptibles, supporting thereby an endless cycle of infection. This obviously does not make sense. So in what follows we will stick to the set (4), as it works for any *γ*_1_ and *γ*_2_.

As in the previous case, Eq. (3), as *t* → ∞, *S, I, R, V*_*S*_ and *V*_*R*_ will approach their asymptotic values *S(*∞*), I (*∞*), R(*∞*), V*_*S*_ (∞) and *V*_*R*_(∞). In general *I (*∞) can be different from zero. These asymptotic values are obtained (except for some special cases when either *γ*_1_ or *γ*_2_ is zero) as the solutions of some simple algebraic equations. Two general cases are distinguished, depending on *η* which may have a critical value

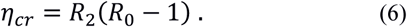

These stationary solutions can be endemic (*η* < *η*_*cr*_) or disease-free (*η ≥ η*_*cr*_). For *η* < *η*_cr_ the asymptotic values read:

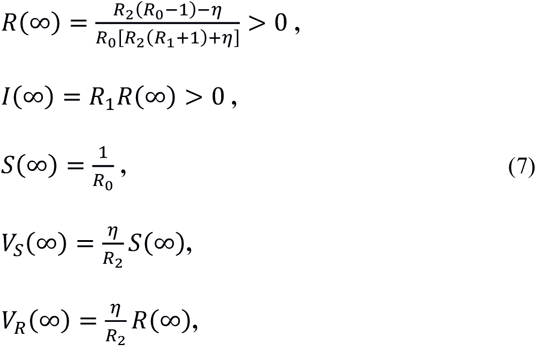

while for *η* ≥ *η*_*cr*_ :

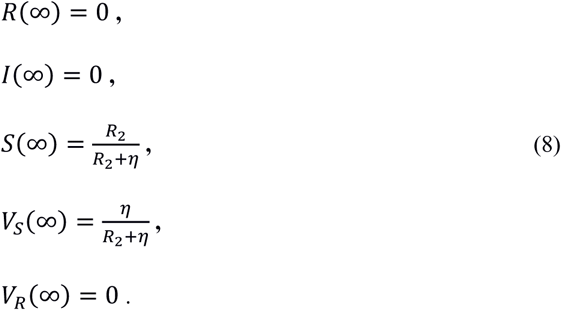

The effective reproduction number asymptotically equals 1 (*η* < *η*_*cr*_) or less than 1 ((*η ≥ η*_*cr*_).

## V. DISCUSSION

In Figs.3-7 we have depicted the most prominent features of the extended model. The spiral-like behavior, as seen in Fig.3 for *η* < *η*_cr_, is due to the fact that an individual leaving a particular compartment, is about to return back after the acquired immunity (triggered by vaccination or by overcoming an infection) expires. Besides this back and forth behavior, the spiral curve never touches the x-axis as the asymptotic value for *I*(∞) is different from zero. Conversely, for *η ≥ η*_cr_, the curve touches the x-axis and shows no evident spiral-like behavior as *I*(∞) = 0. Nevertheless, by closer inspection we see that the blue curve, before hitting the x-axis, starts making a turn but never makes it a full spiral. To make this interesting feature more noticeable, we have depicted in Fig. 4 the blown up versions of these two curves near their endpoints. More importantly, the blue curve (representing a supercritical case) encroaches less into the curves with constant adaptation rate, as compared with the red curve (representing a subcritical case).

**Fig. 3.**
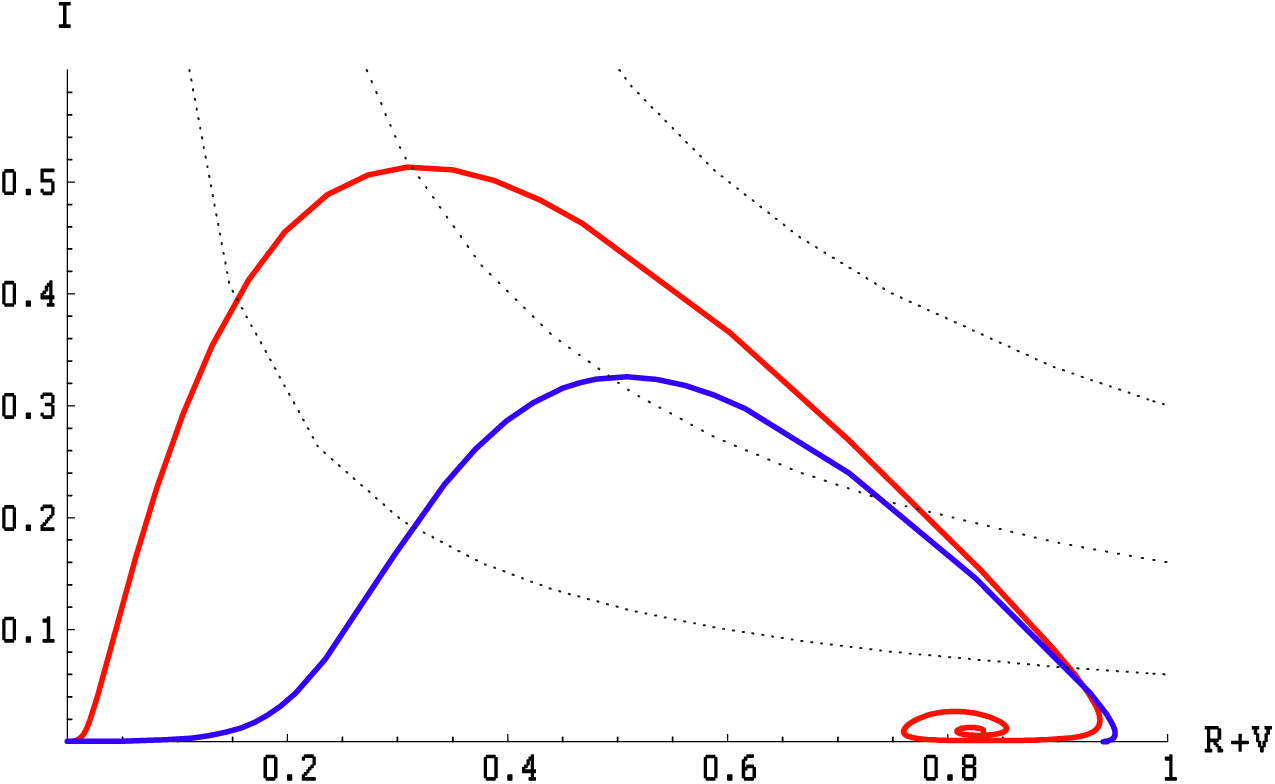
VA-IP diagram for the extended model, Eq. (4). The colored lines show the epidemic dynamics for two different ζ rates: a subcritical ζ=0.004 *d*^*-1*^ (red line) and a supercritical ζ=0.04 *d*^*-1*^ (blue line). In these cases the epidemic starts with *I*(0) = 10^−4^, *R*_o_ = 6, γ=1/4 *d*^*-1*^, γ_1_= γ_2_=1/180 *d*^*-1*^. The critical rate *ζ*_cr_ = *γ* · *η*_cr_ = 0.0278, where *η*_cr_ is given by Eq. (6). The dotted hyperbolas are the same as in Fig.1

**Fig. 4.**
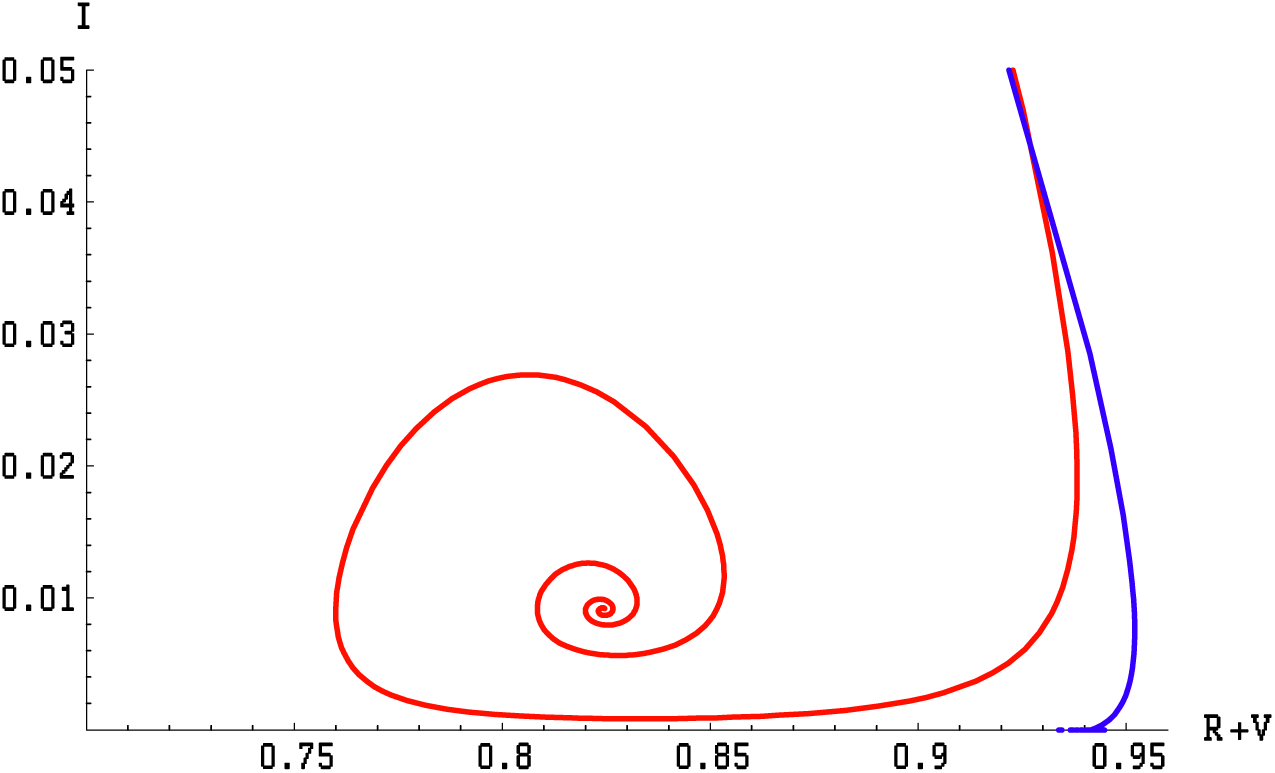
Blown up version of the Fig. 3 in the vicinity of the curves endpoints.

It is clearly seen from Eq. (2) that *EIP* behaves as a straight line in the limit *t* → ∞, and we take its gradient to be a measure of how much of viral adaptation is being transferred to *S(t)*, when stationary states are reached. In Fig. 5 we have depicted the *EIP* for two values of *η*, one subcritical and the other supercritical. In the supercritical case the gradient is zero, meaning not only that a disease is under control but also that viral adaptation stops being transferred soon after the maximum of infection is attained. In the subcritical case instead the gradient is different from zero, signaling not only that a disease is not under control, but also that viral adaptation is being transferred to a susceptible population all the time. In the latter case the number of advantageous mutations grows with time. In Fig. 6 we have shown again the subcritical case retaining the same parameters as in Fig. 5, but now any vaccination is excluded, i. e. *η* = 0 is set. We see a noticeable increase of the gradient in this case, showing that even the low rate vaccination helps in non-proliferation of viral adaptation through a population.

**Fig. 5.**
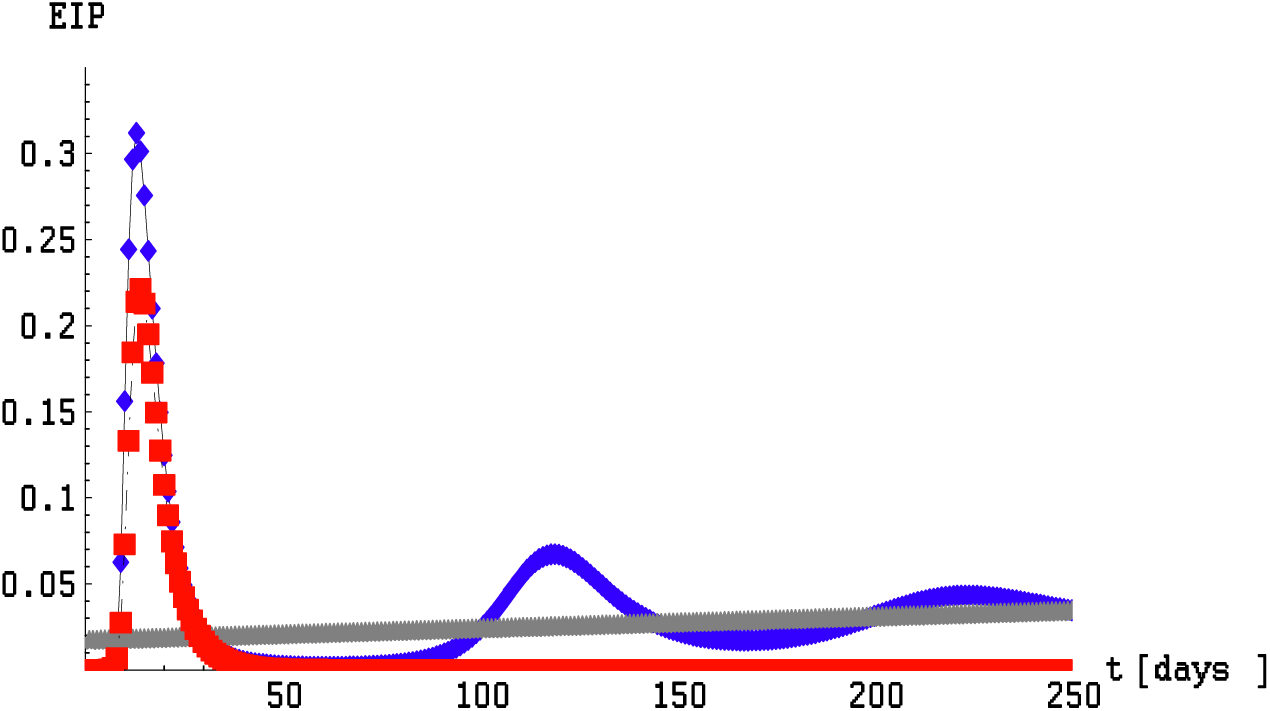
*EIP(t)* function in the extended model, Eq. (4), the parameter space being the same as in Fig.(3). The subcritical *EIP(t)* (ζ=0.004,blue line) behaves as a straight line (gray) with a positive gradient in the limit *t* → ∞. The supercritical *EIP(t)* (ζ=0.04, red line) vanishes as *t* → ∞.

**Fig. 6.**
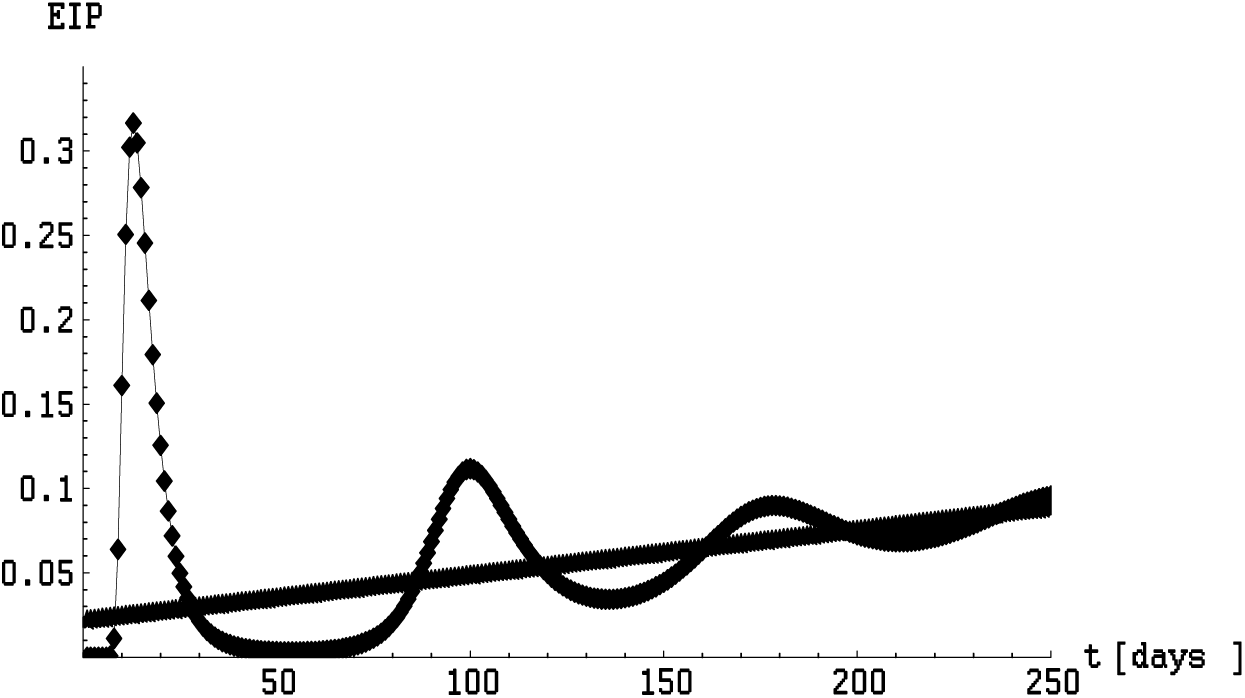
*EIP(t)* with no vaccination (ζ=0), and other parameters being the same as in Fig. 5. *EIP (t)* approaches a straight line as *t* → ∞.

In Fig. 7 we have shown the EIP for two different values of *η*, but now with both values being supercritical. We see a somewhat different pattern of that in Fig. 3 when we increase *η* (analog to increasing *f* there). Apparently the curves do not cross each other, and the potential for transmission of viral mutations shows no tendency to grow at late time for larger *η*. We have also explored large portions of the parameter space from the both sides of *γ*_1_ = *γ*_2_ and found no qualitatively different outcomes.

**Fig. 7.**
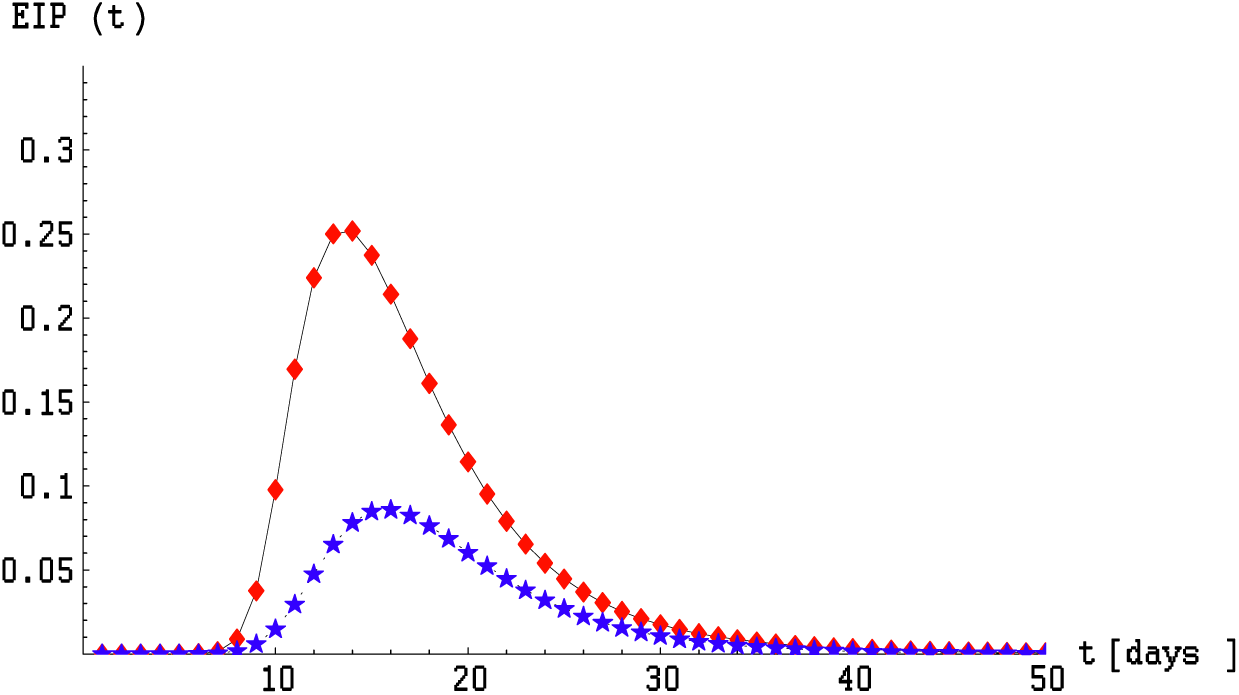
*EIP (t)* for two supercritical cases: ζ=0.03 (red line) and ζ=0.08 (blue line), and the other parameters being the same as in Fig. 5. *EIP*s vanish as *t* → ∞.

Finally, a note about vaccine efficacy (for transmission) in our examples is in order. In our examples we have taken both types of immunity to last for six months, pinpointing the *ζ* critical rate to be 0.0278. This means that almost 3% of the population should acquire vaccine-induced immunity each day. Since the present day vaccines have a pretty low efficacy for the Omicron variant, this means that much more than 3% of the population should be vaccinated each day in order to maintain a disease-free status and prevent further viral mutations. At present this is difficult to achieve for most countries, especially the poorer ones.

## Data Availability

All data produced in the present study are available upon request to the authors

